## Supplementary tables and figures for "Underreporting of SARS-CoV-2 infections during the first wave of the 2020 COVID-19 epidemic in Finland - Bayesian inference based on a series of serological surveys"

### Supplementary tables and figures for "Underreporting of SARS-CoV-2 infections ..." by TA Nieminen et al.

14th February 2023

Table S1: Parameters of the prior distribution in the Estimation model, and the specificities of the screening and confirmation tests.

| Parameter | Description | Value |
|---|---|---|
| $\mu_1$ | Prior expectation for $logit(\pi_1^{(0)})$ | logit(0.05) |
| $\sigma_1$ | Prior standard deviation for $logit(\pi_1^{(0)})$ | 2 |
| $\alpha$ | Shape parameter for gamma prior distribution of $\sigma$ | 2 |
| $\beta$ | Rate parameter for gamma prior distribution of $\sigma$ | 40 |
| $\delta^{Screen}$ | Screening test specificity | 0.9759 |
| $\delta^{Confirmation}$ | Confirmation test specificity | 1 |

Table S2: Influence of choices of prior parameters on the estimation of under-reporting ratio $\Delta(t)$. Shown are posterior means and 95% credible intervals for $\Delta(t)$, based on the confirmation test data, for 9th April 2020 ($t = 0$), 28th May 2020 ($t = 49$) and 2nd July 2020 ($t = 84$), using different values for the parameters $\mu_1$, $\sigma_1$, and $\beta$. The value used for the parameter $\alpha$ was 2.

| Prior parameters | | | Posterior parameters, Confirmation test | | |
|---|---|---|---|---|---|
| $logit(\mu_1)$ | $\sigma_1$ | $\beta$ | $\Delta(t = 0)$ | $\Delta(t = 49)$ | $\Delta(t = 84)$ |
| 0.005 | 2 | 2 | 9.31 (2.571–23.33) | 2.49 (0.355– 7.30) | 4.61 (0.096–23.61) |
| 0.005 | 2 | 20 | 8.17 (3.15–16.05) | 2.48 (0.89– 5.43) | 3.69 (0.78–14.05) |
| 0.005 | 2 | 40 | 8.26 (3.27–15.76) | 2.40 (0.91– 4.89) | 3.04 (0.84– 9.98) |
| 0.005 | 2 | 120 | 8.48 (3.49–15.90) | 2.25 (0.92– 4.31) | 2.22 (0.82– 5.19) |
| 0.005 | 10 | 2 | 9.36 (2.292–24.94) | 2.45 (0.348– 7.13) | 4.53 (0.087–23.05) |
| 0.005 | 10 | 20 | 8.13 (2.97–16.08) | 2.46 (0.84– 5.40) | 3.67 (0.75–13.56) |
| 0.005 | 10 | 40 | 8.28 (3.14–15.76) | 2.41 (0.90– 4.93) | 3.07 (0.81–10.09) |
| 0.005 | 10 | 120 | 8.36 (3.28–15.87) | 2.21 (0.86– 4.31) | 2.17 (0.76– 5.01) |
| 0.050 | 2 | 2 | 11.16 (3.175–28.88) | 2.54 (0.352– 7.32) | 4.61 (0.086–23.02) |
| 0.050 | 2 | 20 | 8.97 (3.54–17.26) | 2.61 (0.94– 5.58) | 3.82 (0.83–14.01) |
| 0.050 | 2 | 40 | 8.95 (3.75–16.50) | 2.56 (1.03– 5.08) | 3.15 (0.93– 9.78) |
| 0.050 | 2 | 120 | 9.22 (3.89–17.02) | 2.43 (1.01– 4.55) | 2.38 (0.89– 5.36) |
| 0.050 | 10 | 2 | 9.47 (2.382–24.46) | 2.49 (0.362– 7.16) | 4.60 (0.088–22.81) |
| 0.050 | 10 | 20 | 8.18 (3.00–16.30) | 2.47 (0.85– 5.37) | 3.68 (0.75–13.80) |
| 0.050 | 10 | 40 | 8.22 (3.20–15.88) | 2.38 (0.90– 4.87) | 2.99 (0.83– 9.64) |
| 0.050 | 10 | 120 | 8.44 (3.37–16.06) | 2.23 (0.88– 4.34) | 2.21 (0.78– 5.21) |

Figure S1: Incidence of COVID-19 cases in the HUS area by age group and language during the first wave of the epidemic in 2020, for Finnish (fi), Swedish (sv), English (en), Russian (ru) and other language groups.

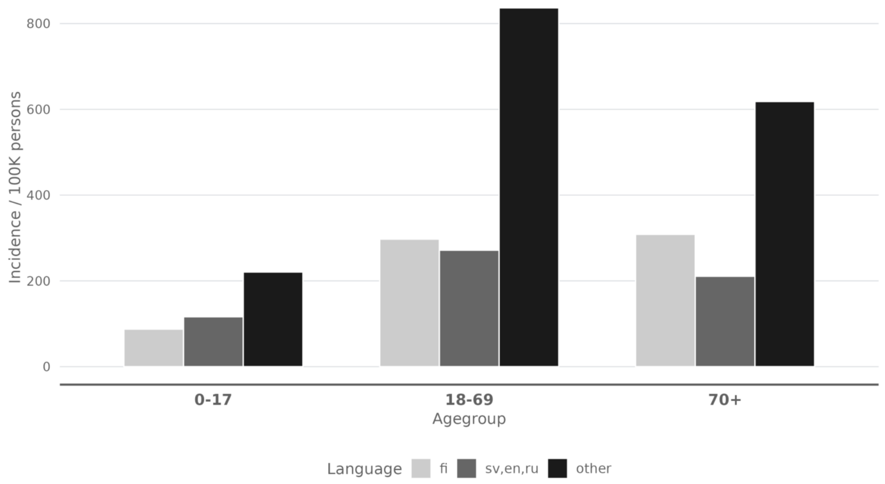

Figure S2: Age distributions of: population in the extended capital region of Finland at the end of 2021 (HUS); COVID-19 cases for the HUS population during the first wave of the COVID-19 epidemic in 2020; the study population, i.e. the target population of the current study (HUS (incl.)); COVID-19 cases from the study population during the first wave of the COVID-19 epidemic in 2020 (FNIDR (incl.)); serological survey participants from the study population during the first wave (Serosurveys).

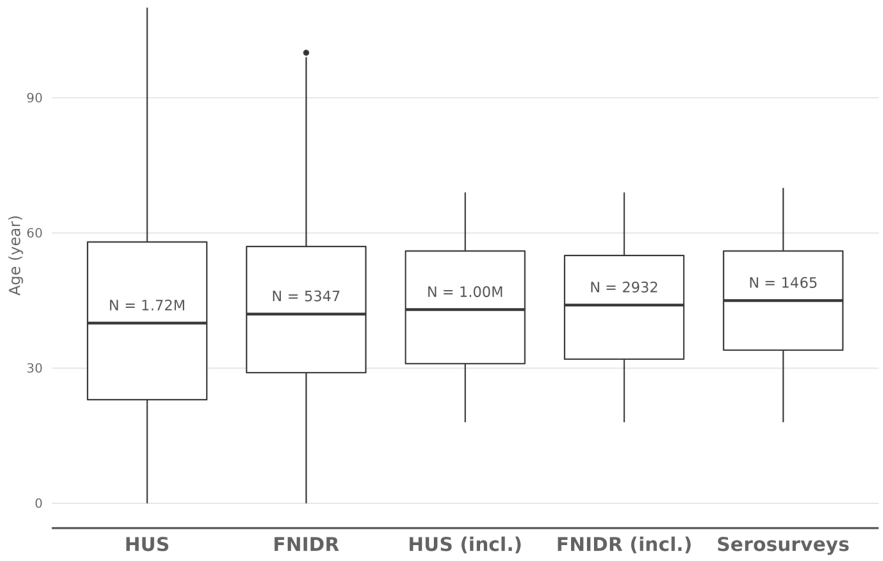

Figure S3: The antibody tests and their performances on the calibration data. The screening test is the result of the IgG antibody test, which may give false positive results. The confirmation test is a combination of the IgG and microneutralization tests (MNT), where the IgG positive samples are tested again with the MNT. After optimizing performance on the calibration data, which includes samples from PCR positive and negative individuals, the sensitivity and specificity of the screening test are 33/33 (100%) and 81/83 (97.59%), respectively, while the sensitivity and specificity of the confirmation test are 33/33 (100%) and 83/83 (100%), respectively.

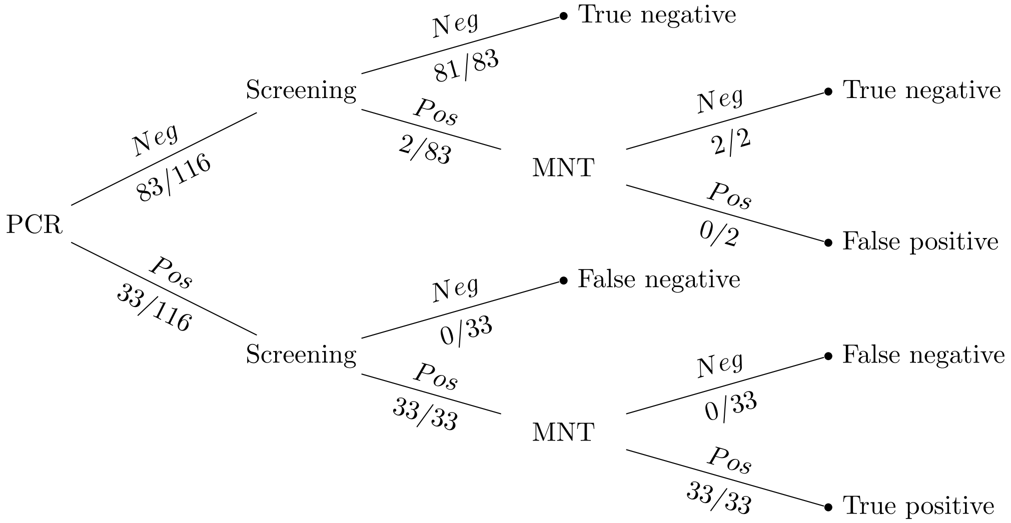

Figure S4: Prior mean, and 2.5% and 97.5% quantiles for each weekly seroprevalence $\pi_w^{(0)}$ in the Estimation model. The estimates were computed based on 40000 samples generated from the prior distribution of $\pi$

.

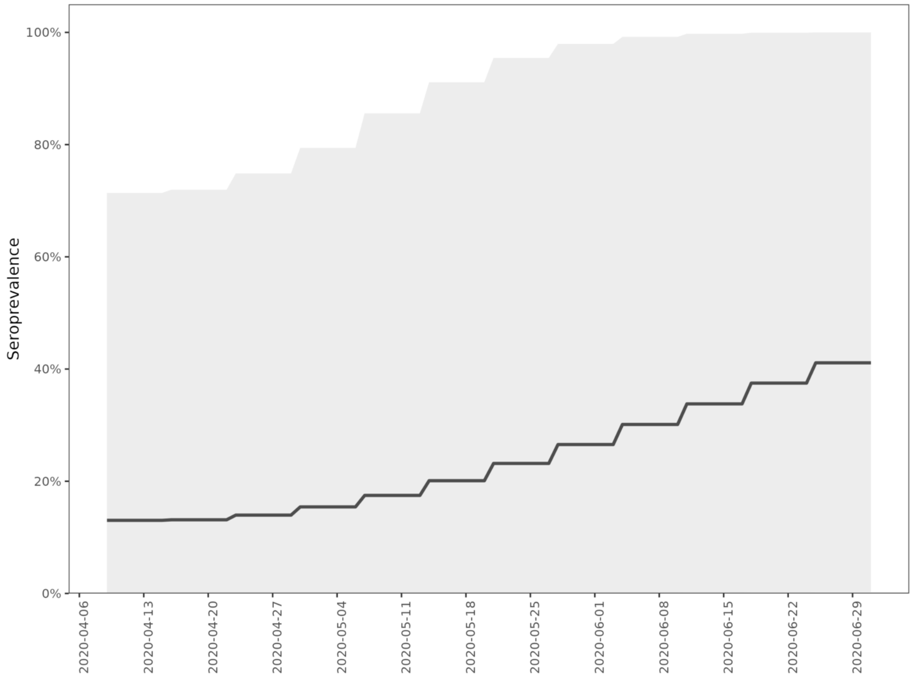


Figure S5: The three images show, starting from the the left: the posterior distribution for $\mu_U$, the posterior distribution for $\sigma_U$, and the posterior predictive distribution for $U$, the time from COVID-19 symptom onset to seroconversion. The distribution for $U$ was obtained by sampling from the lognormal distribution, using samples from the joint posterior distribution for $(\mu_U, \sigma_U)$.

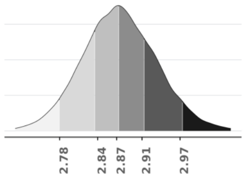


(a) Posterior distribution of $\mu_U$

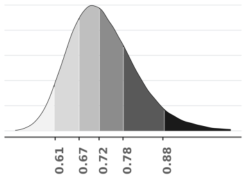


(b) Posterior distribution of $\sigma_U$

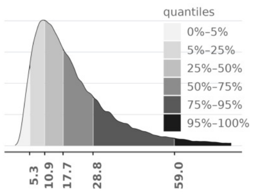


(c) Posterior predictive distribution of $U$.

Figure S6: Prior and posterior distributions for the parameter $\sigma$. Image on the left shows the prior distribution, the middle image shows the posterior distribution based on confirmation test data, and the image on the right shows the posterior distribution based on the screening test data.

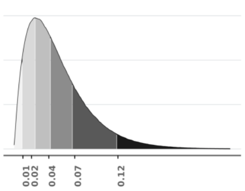


(a) Prior density for $\sigma$.

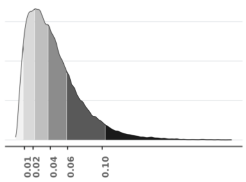


(b) Posterior density for $\sigma$ (Confirmation test).

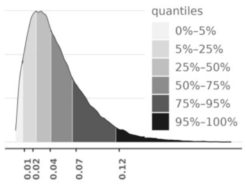


(c) Posterior density for $\sigma$ (Screening test).

Figure S7: Age distribution of COVID-19 cases in the extended capital region of Finland during the first wave of the COVID-19 epidemic in 2020.

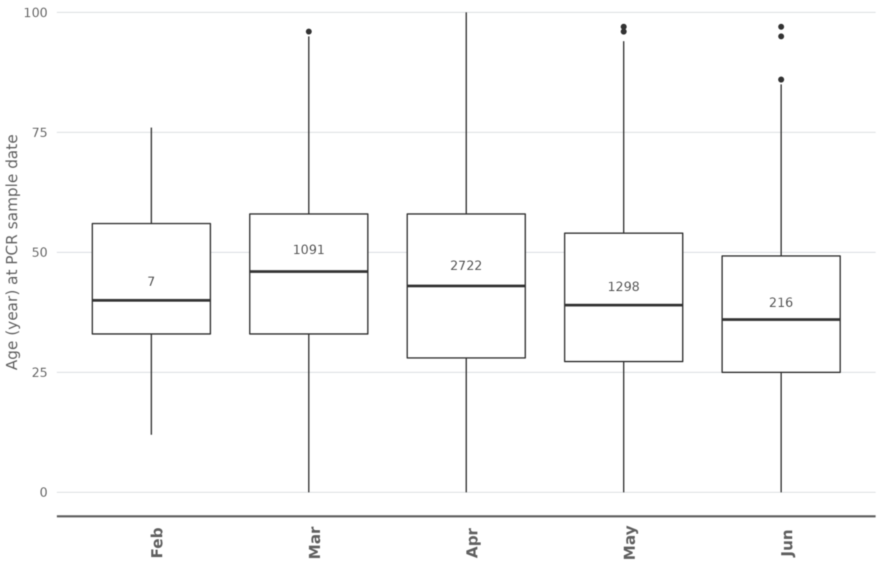
